# Assessing the Quality of Hand-Dug Wells in the Sunyani Municipality, Ghana

**DOI:** 10.1101/2023.08.12.23293919

**Authors:** Henry Ofosu Addo, Antwi Joseph Barimah, Elvis J. Dun-Dery, Fati Ibrahim, Mercy O. Obeng, Christiana Asiamah, Kingsley E. Amegah

## Abstract

**Background:** Hand-dug well water represents one of the main sources of drinking water for many settlements, especially people without access to treated potable water. However, water from such alternative sources can sometimes be polluted and not conducive for human consumption, due to the presence of pathogenic parasitic organisms. This study assessed the physico-chemical and microbial quality of hand-dug wells in the Sunyani Municipality in Ghana.

**Methods:** The study was cross-sectional in nature and results were laboratory-based analyses. The study used Global Positioning System (GPS) tracker to locate and sample hand-dug wells within the Sunyani Municipality. Fifteen (15) households with hand-dug wells were randomly selected from fifteen major communities within the Sunyani Municipality. One (1) sample was collected from each of the 15-household hand-dug wells for the study.

**Results:** Physico-chemical analyses suggest that the temperature of water from all sources ranged between 25.8^**o**^C and 27^0^C whiles phenolphthalein alkalinity of all water samples was 0. But the total alkalinity was between 28 to 116 mg/L. Relatively higher concentrations of nitrate were also observed in samples from all locations and most wells were exposed. However, all samples contained 18.2MPN Index/100ml of faecal coliform compared to 0.0MPN Index/100ml of acceptable national limits.

**Conclusion:** Apart from most samples recording parameters exceeding World Health Organization (WHO) and Ghana Standards Authority recommended levels, majority of wells were exposed and sited around unhygienic environment. Hand-dug well water in the Sunyani Municipality is not completely safe for drinking purposes and requires periodic expert supervision and sample testing.

## Introduction

Water quality constitutes a significant factor in public health especially due to the immense benefits of water to the health and wellbeing of populations. The World Health Organization (WHO) opined that 75% of all diseases affecting developing countries are triggered by drinking polluted water and further observed that about 884 million people across the globe used unimproved water for drinking ^1^.

Groundwater sourced from shallow wells, boreholes and springs remains a major source of water for various uses in Sub-Saharan Africa and other parts of the developing world ^2^. Tapping groundwater from shallow aquifers comes with water quality challenges, especially when these ‘hand-dug wells’ are provided near beneficiary communities and within individual households ^3^. It is worth noting that even the so-called protected deep boreholes are also oftentimes polluted by on-site sanitation facilities. This situation occurs when there is a hydrologic connection between deep aquifers and geologic layers on the earth surface where all on-site sanitation systems are erected ^4^.

The aforementioned situation is more evident in low-income communities and peri-urban areas in Sub-Saharan Africa, where onsite sanitation systems are predominantly used. And sadly, the presence of on-site sanitation systems predisposes groundwater sources to pollution by microbial pathogens ^5^.

Water is the most abundant substance on earth which occupies about 71% of the earth’s total surface ^6^. However, in most developing countries like Ghana, access to quality water and adequate sanitation services is conspicuously lacking. Due to this, much of the urban population solely depend on groundwater aquifer achieved through the construction of hand-dug wells to source water for drinking and other domestic purposes such as for cooking, washing and bathing ^7^. However, in such developing countries, the hand-dug wells are highly susceptible to several sources of pollution ^8,9^.

Contaminants including bacteria, viruses, heavy metals, nitrates and salt have contributed significantly to the pollution of such water sources due to inadequate treatment and disposal of waste from humans and livestock ^10,11^. This therefore, necessitated the need to assess water quality from hand-dug wells within the Sunyani Municipality of Ghana in order to determine their safety for drinking and other relevant uses.

## Methodology

### Study Area and design

The study was done in the Sunyani Municipality which is one of the administrative districts in the Bono Region of Ghana. It lies between Latitudes 20°N and 70.05°N and Longitudes 20.30°W and 20.10°W. Sunyani is the largest settlement in the Bono Region in terms of population and area (Figure 1). The study was cross-sectional in nature and results were laboratory-based analyses. The study used Global Positioning System (GPS) tracker to locate and sample hand-dug wells within the Municipality. The Global Positioning System (GPS) was also used to allocate sample codes for easy identification **(contact corresponding author to request access to these data)**.

### Sampling

The water samples were the main experimental materials. Fifteen (15) households with hand-dug wells were randomly selected from fifteen major communities within the Sunyani Municipality **(contact corresponding author to request access to these data)**. One (1) sample was collected from each of the 15-household hand-dug wells for the study. About 500 ml of water each was taken from the household in the morning between 8 am and 11 am and transported to the laboratory immediately between July and September 2022. Laboratory tests were carried out at the Ghana Water Company laboratory in Sunyani, a well-equipped government laboratory for the Municipality.

### Instrument and Test Procedures for Microbial Quality

Two analytical techniques - Membrane Filtration (MF) and Multiple Tube (MT) were used to determine Total coliform (TC), Faecal coliform (FC), *Escherichia coli* (*E. coli*) and Total heterotrophic (TH) bacteria. To draw water samples from the various houses, the containers used for the collection of samples were first cleaned with 70% ethanol to avoid pre-contamination.

The water samples were collected using hard plastic and screw-capped bottles that have been sterilized to avoid contamination by any physical, chemical or microbial means. The collected hand-dug well water samples were aseptically transferred into sterile containers. Samples for bacteriological analyses were kept in screw-capped bottles that were sterilized in an autoclave for 15 minutes at 121°C. Samples were then transferred to the laboratory for microbial analysis. Aliquots of 100 ml of each sample were separately filtered and each membrane filter placed on appropriate medium for the parameter to be determined. The following microbiological media were used for the membrane filtration analysis: M-Endo Agar for Total coliform bacteria at 37°C for 18-24 hours; MFC Agar for faecal coliform bacteria at 44°C for 18-24 hours; Hichrome Agar for *Escherichia coli* bacteria 37°C for 18-24 hours; and Nutrient Agar for Total heterotrophic bacteria at 37°C for 18-24 hours. For the MTF analysis, 10 ml of sample was dispensed into each of the 10 Lauryl Tryptose tubes containing pre-sterilized broth media.

Bottles were inverted and shaken three to five times to mix contents and to eliminate any air trapped in the inner vial (Durham tube). After that, the tubes were incubated at 35 ± 0.5°C and observed for gas production which was accompanied by a cloudy appearance of the broth at the end of 24 and 48 hours. Positive presumptive results (gas produced) were recorded on the Most Probable Number MPN test data sheet and the corresponding MPN index determined. Following incubation, aliquots from the cultured positive tubes were aseptically streaked on MacConkey agar for Total coliform at 37^0^C.

### Instrument and Test Procedures for Physico-Chemical Quality

The water samples collected were also tested to determine the physico-chemical quality. Different test methods were used to determine pH, Colour, Turbidity, Conductivity, Total dissolved Solids, Hardness, calcium, iron, Nitrite, Fluoride, Arsenic, and manganese. The physico-chemical parameters which were done *in situ* were pH, colour, conductivity, temperature, turbidity and total dissolved solids. The following methods were employed for the various parameters: Electrometric for pH, Conductivity, Total dissolved Solids, Platinum-cobalt for Colour, Nephelometric for Turbidity, Titrimtric for Total hardness, Argentometric for Chloride, Diazotization for Nitrite, Cadmium reduction for Nitrate, Spands for Fluoride, EZ arsenic for Arsenic, and Periodate oxidation for Manganese.

## Results

### Physico-Chemical Test of Hand-Dug Well water

#### Temperature

This study was carried out to assess the suitability of potable water used for domestic activities in fifteen selected suburbs in Sunyani. Water quality parameters such as temperature, pH, electrical conductivity (EC), and total dissolved solids (TDS), were measured. Table 2 is a presentation of laboratory analysis of water samples taken from the different sources. Results are in comparison with national water quality standards specified by the Ghana Water Company Limited. Results indicate that the temperature of water from all sources ranged between 25.8 ^**o**^C and 27^0^C, with Nkwabeng South (926) and Penkwase number 1(924) recording the lowest temperatures of 25.8^0^C. The highest temperature of 27^0^C was recorded at New Town (939). Observable discomfort could be associated with temperatures higher than 36^0^C but temperature levels were within limits for this present study.

**Table 1:** Physico-Chemical Test Results.

| Parameter | Nkwabeng South (926) | South Ridge (919) | Zongo (923) | Penkwase 1 (924) | Berlin top (918) | Estate (922) | Airport (928) | New Town 1 (921) | Nkwabeng North (927) | Baakoniaba (920) | Penkwase 2 (937) | Magazine (938) | Area 3 (930) | Baakoniaba 2 (940) | New Town 2 (939) | Ghana Standard |
| --- | --- | --- | --- | --- | --- | --- | --- | --- | --- | --- | --- | --- | --- | --- | --- | --- |
| Temperature | 25.8 | 25.9 | 26.1 | 25.8 | 26.2 | 26.2 | 26.0 | 26.4 | 25.9 | 26.2 | 26.8 | 26.8 | 26.9 | 26.7 | 27.0 | - |
| pH | 5.39 | 6.09 | 6.30 | 6.30 | 6.40 | 5.83 | 6.36 | 6.07 | 5.81 | 7.18 | 5.86 | 5.71 | 5.33 | 5.87 | 5.51 | 6.5-8.5 |
| Conductivity | 180 | 57.70 | 640 | 210 | 110.10 | 210 | 117.70 | 240 | 180 | 230 | 120.80 | 57.60 | 400 | 141.40 | 290 | - |
| T.D. S | 90 | 28.85 | 320 | 105 | 55.05 | 105 | 58.85 | 120 | 90 | 115 | 60.40 | 28.80 | 200 | 70.70 | 145.00 | 1000 ppm |
| Turbidity | 0.96 | 3.81 | 4.14 | 20.10 | 5.85 | 1.13 | 2.89 | 2.70 | 2.44 | 21.90 | 1.13 | 2.48 | 2.60 | 1.17 | 1.82 | 5 NTU |
| Colour | 25 | 66 | 24 | 54 | 9 | 21 | 38 | 22 | 21 | 62 | 39 | 35 | 23 | 20 | 20 | 0-15 PtCo |
| Total hardness | 38 | 40 | 136 | 74 | 36 | 28 | 62 | 38 | 540 | 54 | 54 | 18 | 58 | 48 | 36 | 500 ppm |
| Alkalinity | 58 | 64 | 100 | 66 | 36 | 28 | 62 | 38 | 28 | 116 | 46 | 50 | 34 | 40 | 28 | - |
| Chloride | 55 | 30 | 162 | 68 | 34 | 78 | 30 | 72 | 68 | 30 | 50 | 32 | 126 | 34 | 82 | 250 ppm |
| Nitrate | 13.64 | 2.64 | 9.24 | 7.04 | 4.84 | 7.48 | 2.64 | 9.24 | 7.04 | 5.28 | 5.72 | 2.20 | 14.08 | 7.04 | 10.12 | 3.0 ppm |
| Sulphate | 1 | 1 | 4 | 8 | 3 | 10 | 1 | 22 | 16 | 8 | 2 | 3 | 0 | 2 | 3 | 250 ppm |
| Iron | 0.04 | 0.12 | 0.04 | 1.63 | 0.08 | 0.04 | 0.24 | 0.03 | 0.09 | 0.59 | 0.06 | 0.02 | 0.04 | 0.00 | 0.03 | 0.3 ppm |
| Phosphate | 0.03 | 0.07 | 0.05 | 0.09 | 0.01 | 0.05 | 0.04 | 0.05 | 0.03 | 0.69 | 12.19 | 0.31 | 0.26 | 0.28 | 0.27 | 0.3 ppm |
| Fluoride | 0.17 | 0.00 | 0.27 | 0.00 | 0.21 | 0.18 | 0.08 | 0.04 | 0.06 | 0.05 | 0.00 | 0.00 | 0.10 | 0.00 | 0.13 | 1.5 ppm |
| Manganese | 0.30 | 0.10 | 0.50 | 0.00 | 0.30 | 0.20 | 1.00 | 0.00 | 0.20 | 0.20 | 0.60 | 0.30 | 0.40 | 0.60 | 0.30 | 0.4 ppm |
| Aluminum | 0.22 | 0.76 | 0.19 | 0.04 | 0.09 | 0.03 | 0.03 | 0.05 | 0.00 | 0.04 | 12.00 | 0.11 | 0.31 | 0.01 | 0.07 | 0.2 ppm |
| Cyanide | 0 | 0 | 0 | 0 | 0 | 0 | 0 | 0 | 0 | 0 | 0 | 0 | 0 | 0 | 0 | 0.07 ppm |
| Lead | 0 | 0 | 0 | 0 | 0 | 0 | 0 | 0 | 0 | 0 | 0 | 0 | 0 | 0 | 0 | - |
| Arsenic | 0 | 0 | 0 | 0 | 0 | 0 | 0 | 0 | 0 | 0 | 0 | 0 | 0 | 0 | 0 | 0.01 ppm |

**Source: Field data, 2022**

**Table 2:** Microbiological Test Result.

| Parameter | Nkwabeng South (926) | South Ridge (919) | Zongo (923) | Penkwase 1 (924) | Berlin top (918) | Estate (922) | Airport (928) | New Town 1 (921) | Nkwabeng North (927) | Baakoniaba (920) | Penkwase 2 (937) | Magazine (938) | Area 3 (930) | Baakoniaba 2 (940) | New Town 2 (939) | Ghana Standard |
| --- | --- | --- | --- | --- | --- | --- | --- | --- | --- | --- | --- | --- | --- | --- | --- | --- |
| *E. coli* | 0 | 0 | 0 | 0 | 0 | 0 | 0 | 0 | 0 | 0 | 0 | 0 | 0 | 0 | 0 | 0.0 MPN Index/100mL |
| Faecal coliform | 18.2 | 18.2 | 18.2 | 18.2 | 18.2 | 18.2 | 18.2 | 18.2 | 18.2 | 18.2 | 18.2 | 18.2 | 18.2 | 18.2 | 18.2 | 0.0 MPN Index/100mL |
| Total viable count | 17 | 21 | 27 | 13 | 7 | 0 | 11 | 33 | 22 | 11 | 0 | 5 | 77 | 44 | 150 | 0-3 CFU |
| *Pseudomonas* | 0 | 0 | 0 | 0 | 0 | 0 | 0 | 0 | 0 | 0 | 0 | 0 | 0 | 0 | 0 | +/- P/A |
| *Clostridium* | 0 | 0 | 0 | 0 | 0 | 0 | 0 | 0 | 0 | 0 | 0 | 0 | 0 | 0 | 0 | +/- P/A |
| *Streptococcus* | 0 | 0 | 0 | 0 | 0 | 0 | 0 | 0 | 0 | 0 | 0 | 0 | 0 | 0 | 0 | +/- P/A |

**Source: Field data, 2022**

#### pH

Considering national guidelines, pH lower than 4 will produce sour taste and higher values above 8.5 give water a bitter taste. From the results presented in Table 2, all but Baakoniaba Number 1(code 920) had water sources with pH levels (7.18) within national standards of 6.5-8.5. Area 3 (code 930) presented the lowest pH value of 5.33. The various indications have suggested that even though pH levels are lower than national standards, water from all fifteen sources is likely to have a normal taste, considering that they are above a minimum pH value of 4 and below a maximum value of 8.5. However, other side effects could be associated with current pH levels apart from taste.

#### Conductivity

Electrical conductivity is a useful tool to evaluate the purity of water, which was minimum at Magazine (code 938 (57.6µ Scm^-1^ at 25 °C)) and maximum at Zongo (code 923) (640µ Scm^-1^ at 25 °C). Relatively higher levels of conductivity were also observed in all other well water samples.

#### TDS

TDS are those solvents which dissolve in water and cannot be separated from water by filtration. They may be chemically organic or inorganic. In the current study, the highest value (320ppm) was recorded at Zongo (923) and the lowest (28.8ppm) at Magazine (938). The difference in values may be due to evaporative loss of water and increase in the concentration of salts present in water. The national standard for dissolvable solids is up to 1000 ppm permissible quantity, and the results indicate that samples from all the fifteen sources were within permissible limits.

#### Turbidity

Considering a national turbidity level of 5 NTU, the analysis revealed that Baakoniaba number 1 (920) had the highest turbidity level (21.9NTU), with Nkwabeng South (926) recording the least value of 0.96 NTU. Other locations that had turbidity levels above national standards included Penkwase number 1 (924), (20.1NTU) and Berlin Top (918), (5.85NTU). Aside these, all other locations presented turbidity levels less than acceptable national specifications for drinking water.

#### Colour

The colourless nature of drinking water is the initial physical assessment of a clean drinking water source. However, except Berlin Top (918) (9pt.co.), water from all other sources contained colour higher than acceptable limits of 0-15pt.co., with South Ridge (919) having the highest (66pt.co).

#### Total Hardness

Hardness has no known adverse effects. Hardness above 500ppm of water is not suitable for domestic use in washing, cleaning, laundry and drinking. The total hardness of water samples ranges from 18ppm at Magazine (938) to 540ppm at Nkwabeng North (927).

#### Alkalinity

The phenolphthalein alkalinity of all water samples is 0. But the total alkalinity for current study was found between 28 to 116 mg/L. According to Environmental Protection Agency, the acceptable limit of total alkalinity of drinking water sample is 500 mg/L CaCO_3_ and maximum desirable limit is 1500 mg/L CaCO_3_. Therefore, all samples contained alkalinity levels within acceptable limits.

#### Chloride

Chloride exists in all natural waters. The concentrations vary very widely and do not pose a health hazard to humans. Principal consideration is in relation to palatability. Chloride values for this study ranged from 30ppm in South Ridge (code 919) to 162ppm at Zongo (code 923). The acceptable desirable limit is 250ppm. Results showed that all samples fell within acceptable national limit for drinking water.

#### Nitrate

Relatively higher concentrations were observed in samples from all locations with Nkwabeng South (926) being the highest (13.64ppm) and Magazine recording the lowest (2.2ppm) Nitrate concentration. South Ridge (code 919) and Airport (code 928) were locations also recording slightly lower (2.64ppm, respectively) nitrate concentrations than national limits for drinking water.

#### Sulphate

Aside the ability of sulphate to give drinking water a bad taste, it can also act as a purgative in humans. The levels of sulphate in the water samples were generally low, ranging between 1ppm in Nkwabeng South (926), South Ridge (code 919), and Airport (code 928) to 22ppm in Newtown (code 939) and 16ppm in Nkwabeng North (code 927). WHO/national limit for sulphate concentration in drinking water is 250ppm. Results from the analysis have indicated that all fifteen samples were within acceptable limits.

#### Iron concentration

Iron is an essential component in human diet. Estimates of the minimum daily intake for iron depend on age, physical status, sex and iron bio-availability. Although iron has got little concern as a health hazard but is still considered as a nuisance in excessive quantities. Samples that were higher than national specifications included those from Penkwase 1 (924), (20.1NTU) (1.63ppm), Baakoniaba number 1 (code 920) (0.59ppm), Airport (code 928) (0.24ppm), and South Ridge (code 919) (0.12ppm). Long-term consumption of drinking water with a high concentration of iron can lead to liver diseases. Aside these, all other samples analyzed were within acceptable limits, as they indicated concentration levels less than 0.3ppm national limit.

#### Phosphate

Phosphate naturally occurs in water and often in high concentrations. The water samples analyzed recorded phosphate concentrations measuring between 12.19ppm from Penkwase number 2 (code 937) samples, 0.69ppm from Baakoniaba number 1 (920) and least concentration of 0.01ppm at Berlin Top (918). Majority of samples analyzed were below national concentration limit of 0.3 ppm.

#### Fluoride

The result suggests that all samples (15 samples) had fluoride concentration less than ppm recommended limit. Zongo (923) recorded the highest concentration of 0.27 ppm, followed by concentration of 0.21ppm from Berlin Top (918), 0.18ppm from Estate (922) and 0.17ppm from Nkwabeng South (926). All other sample concentrations were less than 0.13ppm.

### Trace metals

#### Manganese

Considering a national acceptable limit of 0.4ppm, Airport (1.0ppm), Penkwase Number 2 (0.6ppm), Baakoniaba number 2 (Code 940) (0.6ppm) and Zongo (923) recorded values higher than acceptable limits. On the other hand, all other samples contained manganese lower than 0.4ppm, which is considered acceptable.

#### Aluminum

Aluminum was one metal analyzed for its presence in water samples. The results indicate a trace in samples from South Ridge (0.76ppm), Area 3 (0.31ppm) and Nkwabeng South (0.22ppm) were higher than acceptable limits of 0.2ppm. However, traces of aluminum in other samples were below 0.2ppm limit set for the presence of the metal in drinking water. In analyzing for the presence of other substances, traces of cyanide (national limit of 0.07ppm), lead, and Arsenic (national limit of 0.01ppm) were not recorded in all samples (Table 2).

### Microbiological Test Results

The presence of *E. coli* in water indicates recent faecal contamination and may indicate the possible presence of disease-causing pathogens. From Table 3, analysis of *E. coli* from all fifteen samples had no traces of *E. coli*. However, all samples contained 18.2 MPN Index/100ml of faecal coliform compared to 0.0MPN Index/100ml of acceptable national limits. On Total Viable Count (TVC), only Estate (922) and Penkwase Number 2 (937) recorded no values. Conversely, New Town number 2 (939) had the highest TVC of 150 CFU, followed by Area 3 (77 CFU), Baakoniaba number 2 (44 CFU), and New Town Number 1 (33CFU). Still on TVC, samples from Zongo (923) contained 27 CFU, Nkwabeng North (927) recorded 22 CFU, and 21CFU was recorded in samples from South Ridge. All other samples contained TVC less than 20 CFU (Table 3).

**Table 3:**
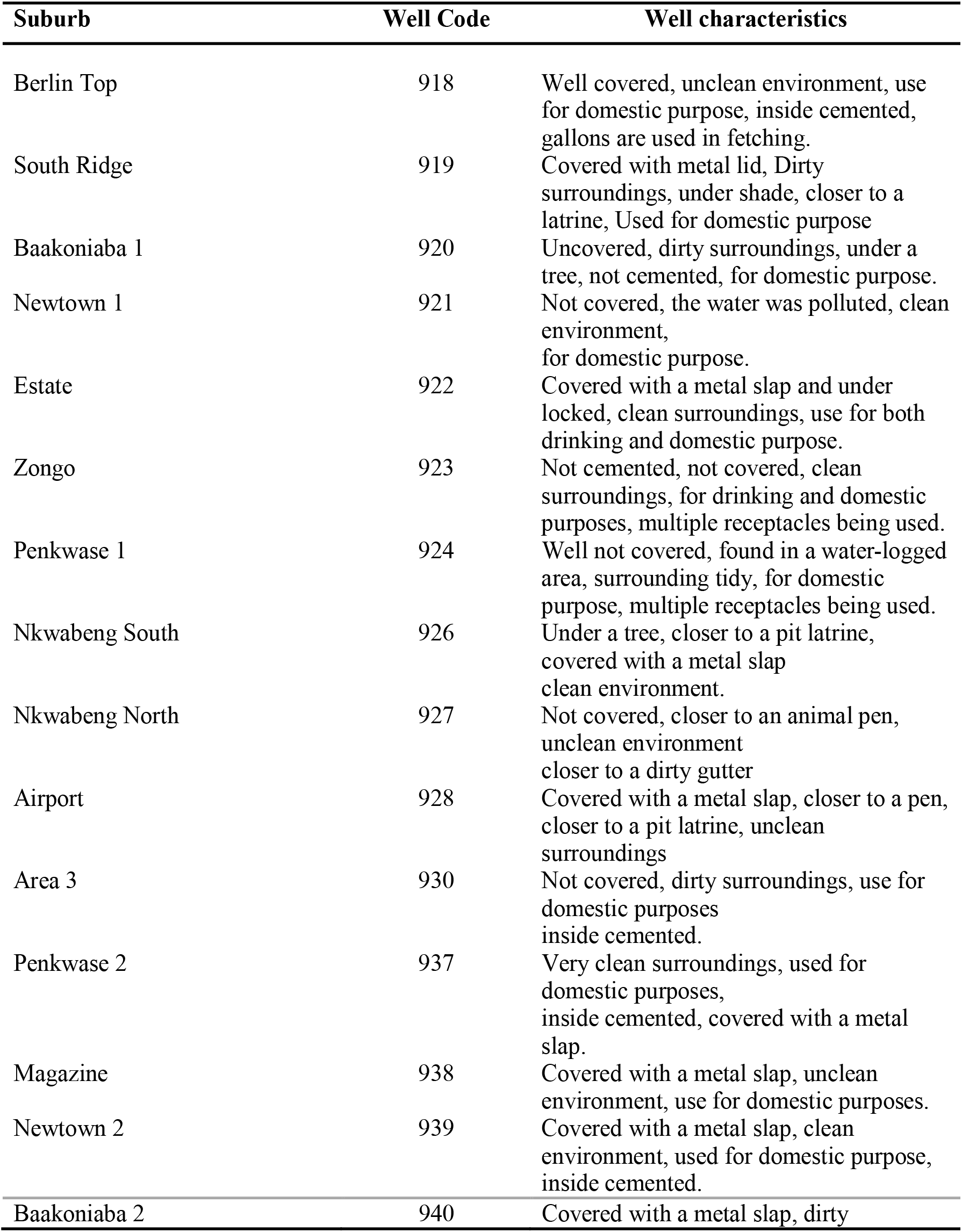
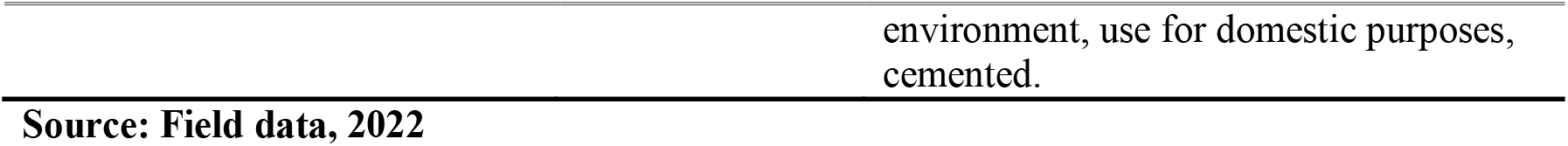
Characteristic Features of Hand-dug Wells in the Sunyani Municipality.

## Discussion

Water of good quality is very essential for sustaining life ^12^. Therefore, a study assessing the quality of water, in this case, hand-dug-wells is very crucial. The general characteristics of the wells used for this study have been listed (Table 4). The results of this study suggest that temperature from all collected samples was within nationally recommended level, as compared to findings ^13^, in Sri Lanka, where variations in water temperature had adverse effects on the quality of water. Variations in water temperature in the two study settings may be due to variations in general atmospheric conditions. Conversely, studies in other parts of Ghana reported a temperature of 40 °C ^14^, far above that reported by the current study. Meanwhile, observable discomforts could be associated with water temperature higher than 36 °C. In similar studies conducted in Northwest of Iran, while researchers reported electrical conductivity (EC) to range between 110 and 1750 μs/cm ^15^, EC values in the current study were lower. Even though the Ghana Ministry of water resources indicated that water colour increases coagulant chemical usage and associated high cost of water treatment ^16^, all water samples collected for this current study had colour above the World Health Organization threshold of 15Pt.Co ^15,17^. Meanwhile, colour in water is known to impede the visibility of physical impurities. Even though nitrate is known to have less effect on the overall quality of drinking water, it is still a public health concern ^18–20^. The presence of high nitrate level in all samples collected for this study was noted. This is slightly different from those reported in Nigeria which ranged between 2.97-40.68 mg/L ^21^. Even though alkalinity reported by this study was within acceptable limits, findings by ^22^ were also within acceptable limits but higher than those reported from current findings. Aside this, several acceptable alkalinity levels have been cited in literature ^23–25^. Nitrate in water is known to pose some level of danger to human health, especially when it is higher than acceptable levels.

In similar studies conducted in Ghana on the quality and health risk of well water ^26^ and water quality of dug-out wells ^14^, the presence of nitrate was reported to be high which agrees with findings of this current study. Contrary to these findings, research from other parts of Ghana reported relatively lower levels of nitrate in well water ^27^. The presence of nitrate can sometimes be attributed to the prolonged use of agricultural fertilizer at close range to hand-dug wells. Manganese is considered an important component of human existence but its presence in drinking water should be measured for health risk purposes. High amounts of manganese in water samples collected from a number of hand-dug wells in sections of Ghana have been recorded ^26^, contrary to findings of our study where the presence of manganese in water samples was lower. Even though the presence of iron is significant to human nutrition ^28^, traces of iron in samples of this study were low. This finding agrees with earlier studies conducted in other parts of Ghana ^10^, but contrary to findings reported by ^29^ where high iron level in well water samples was found. The low presence of iron in water is an indication of water quality since high amount of iron is known to lead to liver diseases and other conditions ^23,28^.

A study in Iran observed variations in levels of fluoride but reported a general level less than 0.5 mg/L ^15^. This, however, is higher than findings of this study as the current research cited only 0.27mg/L as the highest rate of fluoride. It is worth noting that the difference in the study area could have accounted for variations in the levels of fluoride in water samples. Most importantly is the fact that both studies reported traces of fluoride below WHO acceptable levels, since high consumption of fluoride could lead to skeletal and dental fluorosis ^10,30^. In ensuring the wholesomeness of drinking water for human consumption, trace metals are essential elements to investigate.

Laboratory analysis from the current study indicates high presence of faecal coliform in well water samples. This finding concurs with earlier studies ^24,31,32^, where the total faecal coliform count ranged between 3.0 x 102 to 9.3 x104 CFU/ml. Even though both studies reported faecal coliforms higher than the WHO acceptable levels of 0.00 CFU/ml ^33^, the difference may have resulted from the location of the wells, methods of sample analysis or proximity of wells to human settlements. It is also reported in literature that total faecal coliforms in drinking water vary due to seasonal treatment of wells ^34,35^. Other studies have acknowledged the influence of *Streptococcus* in drinking water assessment and ensuring drinking water quality. The current study found no traces of *Streptococcus* in samples ^36,37^.

## Conclusion

Even though majority of samples had parameters within acceptable limits, other indicators show that samples had parameters above limits. Most trace metals were within acceptable limits whiles indicators such as Nitrate and Total Viable Count were above national standards and WHO recommendations. Generally, hand-dug well water is not completely safe in the Sunyani Municipality, considering the exceeding limits of Total Viable Counts, presence of faecal coliform and varying pH levels. Most wells are also left open and sited at unhygienic surroundings. This study has implications for policy direction on the siting of wells for domestic use. It is necessary to periodically conduct well inspection, education on safe water and sample testing to ensure the availability of safe water for users of hand-dug well water. Pathogenic organisms like bacteria if present in drinking water could expose users to various health problems like diarrhea, hence the need for periodic testing to ensure the integrity and safety of the hand-dug well water for consumers.

## Data Availability

All relevant data are within the manuscript

## Ethical Considerations

This study did not involve the use of live participants hence did not require ethical clearance from an institutional review board. However, written informed consent was obtained from owners of all hand-dug wells after the objectives, risk and purpose of the study were explained to them. Voluntary participation was assured and no benefits were given to participants who permitted the collection of samples from their wells.

## Authors’ contribution

**Conceptualization**: Henry Ofosu Addo, Elvis J. Dun-Dery

**Data curation**: Fati Ibrahim, Mercy O. Obeng, Christiana Asiamah

**Formal analysis**: Kingsley E. Amegah

**Investigation**: Henry Ofosu Addo, Antwi Joseph Barimah, Elvis J. Dun-Dery, Kingsley E. Amegah

**Methodology**: Henry Ofosu Addo, Antwi Joseph Barimah, Elvis J. Dun-Dery, Fati Ibrahim

**Writing – original draft**: Henry Ofosu Addo, Antwi Joseph Barimah, Elvis J. Dun-Dery, Kingsley E. Amegah

**Writing – review and editing**: Henry Ofosu Addo, Antwi Joseph Barimah, Elvis J. Dun-Dery, Fati Ibrahim, Mercy O. Obeng, Christiana Asiamah, Kingsley E. Amegah

## Acknowledgement

The authors would like to thank owners of all wells in the study area who permitted the collection of samples for this study and to Research Web Africa for providing technical support.

